# Coping with COVID-19 Pandemic: A Population-Based Study in Bangladesh

**DOI:** 10.1101/2021.03.30.21254632

**Authors:** K M Amran Hossain, Karen Saunders, Mohamed Sakel, Lori Maria Walton, Veena Raigangar, Zakir Uddin, Mohammad Anwar Hossain, Asma Islam, Faruq Ahmed, Rafey Faruqui, Tamanna Tasnim, Shohag Rana, Rubayet Shafin, Md. Shahoriar Ahmed, Md. Obaidul Haque, Md. Feroz Kabir, Mohammad Sohrab Hossain, Iqbal Kabir Jahid, Mst. Hosneara Yasmin, Sonjit Kumar Chakrovorty, Md. Shahadat Hossain, Joty Paul

## Abstract

This study aims to investigate coping strategies used by Bangladeshi citizens during the COVID-19 pandemic.

**Design:** Prospective, cross-sectional survey of adults (N=2001) living in Bangladesh.

**Methods:** Participants were interviewed for socio-demographic data and completed the Bengali translated Brief-COPE Inventory. Statistical data analysis was conducted using SPSS (Version 20).

**Results:** Participants (N=2001), aged 18 to 86 years, were recruited from eight administrative divisions within Bangladesh (mean age 31.85±14.2 years). Male to female participant ratio was 53.4% (n=1074) to 46.6% (n=927). Higher scores were reported for *approach* coping styles (29.83±8.9), with lower scores reported for *avoidant* coping styles (20.83 ± 6.05). *Humor* coping scores were reported at 2.68±1.3 and *religion* coping scores at 5.64±1.8. Both men and women showed similar coping styles. Multivariate analysis found a significant relationship between male gender and both *humor* and *avoidant* coping (p <.01). Male gender was found to be inversely related to both *religion* and *approach* coping (p <.01). Marital status and education were significantly related to all coping style domains (p<.01). Occupation was significantly related to *approach* coping (p <.01). Rural and urban locations differed significantly in participant coping styles (p <.01). Factor analysis revealed two cluster groups (Factor 1 and 2) comprised of unique combinations from all coping style domains.

**Conclusion:** Participants in this study coped with the COVID-19 pandemic by utilizing a combination of coping strategies. Factor 1 revealed both *avoidant* and *approach* coping strategies and Factor 2 revealed a combination of *humor* and *avoidant* coping strategies. Overall, a higher utilization of *approach* coping strategies was reported, which has previously been associated with better physical and mental health outcomes. *Religion* was found to be a coping strategy for all participants. Future research may focus on understanding resilience in vulnerable populations, including people with disability or with migrant or refugee status in Bangladesh.

## INTRODUCTION

The arrival of the COVID-19 pandemic in Bangladesh, in March 2020, adversely impacted the health and everyday lives of Bangladeshi citizens in a variety of stressful ways.^[1]^ Bangladesh is currently ranked 8th in the world in terms of estimated size of population and 6th by density^[2]^ and, as a country, has suffered from a history of natural disasters including flooding, drought, and cyclones.^[3]^ The new coronavirus, SARS-CoV-2, has severely impacted communities that were already under substantial socio-economic pressures prior to the pandemic. The negative impact of COVID-19 on socio-economic resources, in addition to the high prevalence of illness and deaths from COVID-19 infection, have created new and complex challenges and vulnerabilities for the people of Bangladesh, including specific health concerns, food security, and nutrition.^[4]^

The first positive case of COVID-19 diagnosed was reported on March 8, 2020.^[5]^ Since then, 562,752 cases of positive COVID-19 have been reported with 8,608 deaths and 515,989 individuals who have recovered and survived.^[6]^ The actual number of COVID-19 related deaths will be is much higher than the reported number. The authors believe that this is likely due to the significant under-reporting of COVID-19 related deaths in Bangladesh, which cannot be accounted for at this time. The World Health Organization (WHO) dashboard shows a spike of cases from May to October 2020 (First wave); from December 2020 to January 2021 (Second wave) ^[7]^ and in March 2021, a rise in the number of newly diagnosed cases, where treatment within an Intensive Care Unit has been required and reported. ^[8]^ A cross-sectional sample survey of the general population, from March 2020 to December 2020, examined the knowledge, attitudes and behaviors toward COVID-19 across all provinces in Bangladesh and reported a high prevalence of positive, preventive health behaviors, including the wearing of face masks in public places.^[9]^ It was also evident that “fear” and “knowledge of COVID-19” was strongly associated with the practice of such preventive health behaviors. Since then, it is possible that a considerable change in coping strategies, related to the pandemic, may have taken place in Bangladesh. Widespread challenges, including mental health for frontline healthcare workers and the general population in Bangladesh during the Lockdown periods, continue to create further complications on an already overburdened health care system.^[10]^

Prior to the COVID-19 pandemic, Bangladesh had already reported a severe burden on its healthcare sector, including issues related to inefficiency, scarcity of human resources, and corruption. ^[11]^ The COVID-19 pandemic added an additional burden to an already overburdened healthcare system. ^[11]^ It quickly became apparent that the pandemic was precipitating an acute mental health crisis with profound impacts in people of all age groups. ^[10]^ During the first wave, COVID-19 increased fear, ^[9]^ which necessitated multiple approaches of coping. Many people experienced considerable stress because of fear and anxiety, arising from the threat of COVID-19 disease during the first wave. ^[12]^ In Bangladesh, the prevalence of COVID-19 related depression and suicidal ideation, during this time, was reported at 33% and 5%, respectively. The risk factors for depression and suicidal ideation include being young, female, smoking and/or having comorbid disease. ^[13]^ The imposed lockdown intervention, self-isolation, and quarantine measures only intensified mental health challenges across the country, as the majority of people were placed in survival mode and left without any form of societal and/or financial support. ^[14]^ This resulted in an increased report of loneliness, boredom, and anger during lockdown and may have contributed toward the adoption of negative approach coping styles.

Historically, humans have developed various coping strategies to survive disasters. ^[15]^However, most people exhibit poor mental coping strategies over a prolonged period of time. ^[16]^ Coping strategies are defined as “the activities one does to tolerate or decrease mental strain”. ^[17]^ The Brief-COPE, ^[18]^ a primary tool for measurement of effective and ineffective ways to cope with stressful situations, introduces four key coping styles seen in patients with severe diagnosis, including: (1) Approach Coping Strategy (APC), (2) Avoidant Coping Strategy (AVC), (3) Humor (H), and (4) Religion (R). In relationship to disease, coping strategies are understood to be either “positive” or “negative” or a combination of both traits. Positive coping, as seen in APC strategy is associated with higher adaptation to adversity, better health outcomes, and more stable emotional response to disease. In contrast, negative coping, as seen in AVC strategy is associated with negative attitudes toward the disease, poor physical health and less effective mental health management. ^[18]^ Other categories, such as humor and religion, may include both positive and negative coping approach attributes and are independent of either approach.

The Centers for Disease Control and Prevention (CDC) in the United States advised people to learn how to cope with COVID-19 related stress in more healthy ways in order to develop resilience. ^[19]^ The World Health Organization (WHO) has prioritized positive coping as a strategy to deal with fear, anxiety, sadness, worry, depression, insomnia, physical illness, substance use and other mental health issues. ^[20]^ Global concerns have been reported regarding the utilization of negative coping approaches, such as substance use, which are known to be related to a cascade of negative health outcomes at the societal, community, family and individual levels. Negative coping strategies are, also, frequently linked to an over exposure to more negative information about COVID-19. Hence, those with negative coping strategies tend to ignore important objective information crucial to prevention. ^[21-23]^ There is an emphasis in the global media on sensationalist reporting of negative news, related to the number of people infected and/or deaths from COVID-19. This, in turn, may increase the negative coping strategies related to the perceived risk of COVID-19 in the local population. ^[24]^ Negative coping strategies, though common for some age groups, are not the primary coping strategies reported as used by older adults. Older adults, a vulnerable population at higher risk of COVID-19, have reported more positive approaches to coping than younger counterparts, using resilience and life experience as a guide through the pandemic. ^[25]^ Factors, such as *secondary and higher education*, ^[26]^ have, also, been linked to positive coping strategies. However, other factors, such as *female gender* and *single or separated marital status* have been found to be associated with poor coping strategies. ^[27]^ Furthermore, at the individual level, *personality* may also influence the choice of coping styles, with specific styles of self-blame, distraction, venting and anger associated with poor mental health outcomes. ^[28]^ Religion is a common coping mechanism for people during times of crisis, which may lead to an over dependency on religious institutions for perceived truths and less belief of scientific evidence, especially when there are discrepancies in the public health messages given by different governments and religions. However, this may also provide an opportunity for religious and private institutions to take an active role in supporting positive and effective public health messages for their local communities, and provide another layer of systemic support for public mental and physical health preventive measures. ^[9]^

Coping strategies that are associated with better mental health include humor, acceptance and positive reframing. ^[29]^ Encouraging the adoption of these strategies during the pandemic, may provide one mechanism of facilitating the mental health of individuals and families. The widespread societal behavioral change, across the world, in response to the pandemic has shown an ongoing, pervasive shift in mental health response mechanisms to COVID-19. ^[30]^ In Low to Middle Income Countries (LMIC), including Bangladesh, mental health may not be a priority on the political agenda nor a central focus for funding, both internally and internationally. However, understanding mental health coping strategies may be one mechanism to understanding the differing adherence to public health measures and create a path for future success in the pandemic response. Therefore, the purpose of this research was to explore the range of coping styles utilized by a diverse sample of adult subjects, living across 8 regions in Bangladesh, and to gain insight into the predominant coping strategies and behaviors for future planning of educational initiatives, health policies, and the promotion of better mental health for Bangladeshi men and women. This research is also a foundation for future research on resilience programs to support physical and mental health throughout the lifetime.

## METHODOLOGY

### Study Design

This was a prospective, cross-sectional study of adult men and women, across 8 regions in Bangladesh.

#### Participants

Adult participants, ages 18 and above, were invited to take part in the “Brief-COPE Inventory”, ^[18]^ from October 2020 to January 2021. Participants were recruited via open call and personal communication, and were approached for informed consent for face to face interviews with face to face completion of the Brief-COPE ^[31]^ (S1 File), in addition to providing basic demographic information for the study. The aim of this research was to gain insight and understanding into the main coping strategies, and subsequent coping styles, participants employed during the COVID-19 pandemic in Bangladesh.

The total population of Bangladesh is estimated to be 165,760,741 ^[2]^ and by the end of January 2020, 535,139 cases of positive COVID-19 infection were reported, ^[32]^ diagnosed by real-time polymerase chain reaction test (RT-PCR). Respondent cluster estimates were divided into 8 administrative divisions of the country and the sample size was calculated using “EPI INFO” version 7.4.2.0 developed by the CDC in the US. The sample size calculation was estimated to be 1088, with 99.9% confidence interval, 50% of expected frequency, 5% margin of error, and 1.0 design effect. For 8 geographical clusters, the sample size of 136 for each cluster was used.

### Ethical Permission

Ethical permission was obtained from the Institutional Review Board (IRB) at the Institute of Physiotherapy, Rehabilitation, and Research of Bangladesh Physiotherapy Association on the June 4, 2020 (BPA-IPRR/IRB/04/06/2020/068). Trial registration was obtained prospectively from the Clinical Trial Registry of India (CTRI/2020/10/028196) on October 1, 2020. All participants were given verbal briefs on the study objectives and voluntary written consent was obtained prior to data collection. Participants were assured of confidentiality, ethics and privacy issues and were maintained throughout the study, according to the principles of the Helsinki Declaration ^[33]^ (S 2 File).

### Study procedure

Data collection was undertaken, voluntarily, by 138 undergraduate students from Bangladesh Health Professions Institute (BHPI). After gaining ethical permission, the data collectors were trained, online, regarding the study objectives, ethics, Brief-COPE, and the process of data collection. After training, feedback and evaluation from the data collectors was used to refine the process. The data collectors reside in 55 of the 64 districts within the 8 administrative divisions of Bangladesh that were selected. Data collection was conducted through face to face interviews within these 8 regions, with all data collectors adhering to COVID-19 preventive precautions using personal protective equipment (PPE) and social distancing measures. The respondents completed the Bengali translated version of the Brief-COPE questionnaire, with support from the data collectors as requested by the respondents. A hard paper copy of each completed consent form and questionnaire was scanned to a password protected email for trial record keeping by the primary investigator. A small-scale pilot study was conducted to examine the applicability and feasibility of the larger scale study prior to implementation. All relevant safety and preventive health measures were implemented and maintained by the data collection team throughout the study.

### Questionnaire

The questionnaire consisted of two parts. The first aimed to gather the socio-demographic characteristics of the participants with six questions related to gender, age, marital status, education, occupation, and place of residence. This part also included four questions related to COVID-19 presence or absence of symptoms, as per the Ministry of Health and Family Welfare in Bangladesh [34], COVID-19 positive status, per a RT-PCR test, and COVID-19 positive status of anyone close to them. Reported symptoms included fever, dyspnea (shortness of breath), pneumonia, cough, sore throat, muscle weakness, rhinitis, abdominal pain, vomiting, diarrhea, muscle pain, anosmia, ageusia, sicca syndrome, and fatigue. The second part of the questionnaire focused on coping strategies as identified in the Brief COPE questionnaire ^[31]^ (S1 File). The Brief-COPE Inventory is a 28-item questionnaire, ^[31]^ developed from the original 60 item Coping to Orientation of Problems Experienced (COPE) Inventory scale. ^[35]^ Each item on the Brief-COPE has 4 possible response categories, ranging from 1 to 4. In the inventory, “1” corresponds to *I haven’t been doing this at all*; “2”, *A little bit*; “3”, *A medium amount*, and 4, *I’ve been doing this a lot*. All items except for Humor and Religion are mapped onto the 2 theoretical constructs of Avoidant Coping (AVC) and Approach Coping (APC) and results are calculated using the raw scores for participants in these domains. ^[35]^ In addition, the instrument identifies 14 subscales within it, each linked to 2 items, as follows: “Self-distraction” (SD) (Items 1 and 19); “Active coping” (AC) (Items 2 and 7); “Denial” (D) (Items 3 and 8); “Substance use” (SU) (Items 4 and 11); “Seeking emotional support” (SES) (Items 5 and 15); “Seeking informational support” (SIS) (Items 10 and 23); “Behavioral disengagement” (BD) (Items 6 and 16); “Venting” (V) (Items 9 and 21); “Positive reframing” (PR) (Items 12 and 17); “Planning” (P) (Items 14 and 25); “Humor” (H) (Items 18 and 28); “Acceptance” (A) (Items 20 and 24); “Religion” (R) (Items 22 and 27) and “Self-blame” (SB) (Items 13 and 26)”. AVC is linked to the 6 subscales: D, SU, V, BD, SD and SB, whilst APC is linked to the 6 subscales: AC, PR, P, A, SES and SIS. In terms of more broadly defined coping strategies, the subscales of A, SES, H, PR and R have been described as “emotion focused”. AC, SIS and P have been described as “problem-focused” strategies and BD, D, SD, SB, SU and V have been described as “dysfunctional” strategies. ^[35]^ Thus, the Brief COPE questionnaire enables individuals to express hypothetical responses in terms of coping styles to a potential natural disaster event, such as the COVID-19 pandemic. ^[35]^ In so doing, a spectrum profile across the 2 constructs of AVC and APC can be obtained, along with gaining insight and understanding of the mixture of strategies employed by individuals, when confronted by a stressful life event. Regarding interpretation, AVC is associated with a worse physical health status for those with medical conditions and has also been shown to be less effective at controlling anxiety when compared to AP. ^[35]^ Whereas, APC is associated with an individual person being able to consciously make a more constructive response to adversity, which includes positive adaptive practical adjustments, which can promote better health and emotional responses.

### Reliability and validity of the Brief COPE

This questionnaire has been used in a number of sample population studies related to the experience of stressors in a health-related context. ^[36-41]^ Within these studies, the scale has demonstrated an acceptable level of internal consistency (alpha = 0.70) and good convergent validity regarding use with depression; with marginal test-retest reliability (test-retest = 0.6). Confirmatory factor analysis (CFA) also demonstrated a good model fit and acceptable reliability (Cronbach alpha reliability coefficient = 0.61) of the adapted scale. For this research, the Brief-COPE was translated forward into Bengali and then back translated into English by two bilingual British and Bangladeshi researchers. Content validity was later reviewed and determined by a bilingual epidemiologist and a psychiatrist. In this study, the Cronbach alpha reliability coefficient of the Bangladeshi translated version of the Brief-COPE was determined to be .91.

### Statistical Testing

All data was collated within an excel workbook spreadsheet and analyzed using statistical software to obtain the relevant statistical analyses. We used the Statistical Package software for Social Science, SPSS Version 20.0, for this study. Socio-demographic data was analyzed using descriptive statistics in accordance with the nature of the data. After data collection was completed from 2028 participants, further data audit found 2001 data to be eligible for data analysis. The average and standard deviation values for the numerical data were calculated. Normal distribution and skewness and kurtosis value were evaluated using the Kolmogorov-Smirnov test and Shapiro-Wilk test. The categorical variables related to socio-demographics were considered utilizing non-parametric statistics and scores for the Brief-COPE Inventory were calculated as parametric data, per the normality test results. Relationships between the categorical and the parametric data and between the two data categories were primarily determined by an independent t-test for two variables and chi-square test for more than 2 variables (Table 1). Further binary logistic regression was performed for categorical data with alternating respondents and covariates (Table 2). The relationship between parametric data was determined by Pearson Correlation Coefficient (PCC). An Exploratory Factor Analysis was completed for all the categorical domains in the Brief-COPE Inventory. We conducted a Principal Component Analysis with a Varimax rotation in order to extract the maximum variance from the data set with each component. The correlation matrix was determined by using the Kaiser-Meyer-Olkin (KMO) Test for Sampling Adequacy and further analysis was undertaken if the value obtained was >.50 and Bartlett’s Test of Sphericity tested for significance. Any individual variables with a correlation matrix >.40 were considered to be contributing factors. The correlation matrix was calculated and considered to be of value if the figure obtained was more than or equal to .80. In addition, the minimal percentile eigenvalue cut off rule was determined to be 1.104. The minimum threshold for each factor to be considered was set as .40 (Figure 3). The alpha value was set as p<.05.

**Table 1:** Socio-demographic distribution and relationship with the coping domain

| Variables | Number of participants, n (%) | Avoidance mean± SD | t/Chi | p-value (2tailed) | Approach | t/Chi | p-value (2tailed) | Humor | t/Chi | p-value (2tailed) | Religion | t/Chi | p-value (2tailed) |
| --- | --- | --- | --- | --- | --- | --- | --- | --- | --- | --- | --- | --- | --- |
| **Gender** |  |  |  |  |  |  | .114 |  | 4.7<sup>a</sup> | .001 |  | -1.36<sup>a</sup> | .174 |
| Male | 1074 (53.4) | 21.39±6.0 | 4.80<sup>a</sup> | .001 | 30.13±8.7 |  |  | 2.82±1.4 |  |  | 5.58±1.9 |  |  |
| Female | 927 (46.6) | 20.18±5.9 |  |  | 29.49±9.1 | 1.579<sup>a</sup> |  | 2.53±1.9 |  |  | 5.70±1.8 |  |  |
| **Age** |  |  |  |  |  |  |  |  |  |  |  |  |  |
| 18-35 | 1406 (70.3) |  | 85.5<sup>b</sup> | .831 |  | 120.4<sup>b</sup> | .194 |  | 25.5<sup>b</sup> | .111 |  | 27.8<sup>b</sup> | .065 |
| 36-60 | 496 (24.8) |  |  |  |  |  |  |  |  |  |  |  |  |
| 61-75 | 76 (3.8) |  |  |  |  |  |  |  |  |  |  |  |  |
| 76-90 | 23 (1.1) |  |  |  |  |  |  |  |  |  |  |  |  |
| **Marital status** |  |  |  |  |  |  |  |  |  |  |  |  |  |
| Single | 1043 (52.1) |  | 296.0<sup>b</sup> | .001 |  | 155.2<sup>b</sup> | .002 |  | 57.5<sup>b</sup> | .001 |  | 30.5<sup>b</sup> | .003 |
| Married | 889(44.4) |  |  |  |  |  |  |  |  |  |  |  |  |
| Widowed | 48 (2.4) |  |  |  |  |  |  |  |  |  |  |  |  |
| Divorcee | 21 (1) |  |  |  |  |  |  |  |  |  |  |  |  |
| **Education** |  |  |  |  |  |  |  |  |  |  |  |  |  |
| No formal education | 81 (4) |  |  |  |  |  |  |  |  |  |  |  |  |
| Primary education | 190(9.5) |  |  |  |  |  |  |  |  |  |  |  |  |
| Secondary education | 325(16.2) |  | 337.6<sup>b</sup> | .001 |  | 296.1<sup>b</sup> | .001 |  | 92.0<sup>b</sup> | .001 |  | 90.26<sup>b</sup> | .001 |
| Higher secondary | 713 (35.6) |  |  |  |  |  |  |  |  |  |  |  |  |
| Bachelor | 508(25.4) |  |  |  |  |  |  |  |  |  |  |  |  |
| Masters | 170 (8.5) |  |  |  |  |  |  |  |  |  |  |  |  |
| Ph. D. | 14 (.7) |  |  |  |  |  |  |  |  |  |  |  |  |
| **Occupation** |  |  |  |  |  |  |  |  |  |  |  |  |  |
| Government job | 117 (5.8) |  |  |  |  |  |  |  |  |  |  |  |  |
| Private job | 199 (9.9) |  |  |  |  |  |  |  |  |  |  |  |  |
| Farmer | 40 (2) |  | 305.2<sup>b</sup> | .041 |  | 365.9<sup>b</sup> | .001 |  | 77.4<sup>b</sup> | .005 |  | 75.5<sup>b</sup> | .005 |
| Business | 175 (8.7) |  |  |  |  |  |  |  |  |  |  |  |  |
| Student | 966(48.3) |  |  |  |  |  |  |  |  |  |  |  |  |
| Retired | 52(2.6) |  |  |  |  |  |  |  |  |  |  |  |  |
| Housewife | 359 (17.9) |  |  |  |  |  |  |  |  |  |  |  |  |
| Unemployed | 34 (1.7) |  |  |  |  |  |  |  |  |  |  |  |  |
| Others | 59 (2.9) |  |  |  |  |  |  |  |  |  |  |  |  |
| **Living area** |  |  | - |  |  |  |  |  |  |  |  |  |  |
| Rural | 636 (31.8) | 21.37±6 | 2.732<sup>a</sup> |  | 30.98±8.2 | - |  | 2.78±1.43 | - |  | 5.77±1.85 | - |  |
| Urban | 1365 (68.2) | 20.57±6 |  | .006 | 29.30±9.2 | 3.944<sup>a</sup> | .001 | 2.64±1.31 | 2.271<sup>a</sup> | .023 | 5.58±1.91 | 2.148<sup>a</sup> | .032 |
| **Tested COVID** |  |  |  |  |  |  |  |  |  |  |  |  |  |
| Yes | 70 (3.5) | 21.93±5.33 | 1.562<sup>a</sup> | .121 | 33.64±7.13 | 2.678<sup>a</sup> | .007 | 2.76±1.45 | 0.461<sup>a</sup> | .645 | 6.17±1.45 | 2.403<sup>a</sup> | .016 |
| No | 1931 (96.5) | 20.79±6.07 |  |  | 29.72±8.98 |  |  | 2.68±1.35 |  |  | 5.62±1.90 |  |  |
| **COVID** |  |  |  |  |  |  |  |  |  |  |  |  |  |
| Symptoms Present | 185 (9.2) | 22.23±4.52 | 3.314<sup>a</sup> | .001 | 34.15±6.42 | 6.962<sup>a</sup> | .001 | 2.63±1.23 | -.595<sup>a</sup> | .552 | 6.23±1.54 | 5.168<sup>a</sup> | 0.001 |
| No | 1816 (92.8) | 20.68±6.17 |  |  | 29.39±9.05 |  |  | 2.69±1.37 |  |  | 5.57±1.91 |  |  |
| **COVID** |  |  |  |  |  |  |  |  |  |  |  |  |  |
| **Positive** | 87 (4.3) | 21.91±4.93 | 1.706<sup>a</sup> | .088 | 36.26±6.94 | 3.668<sup>a</sup> | .001 | 2.60±1.13 | -.603<sup>a</sup> | .564 | 6.25±1.66 | 3.104<sup>a</sup> | .002 |
| Not tested or Negative | 1914 (95.7) | 20.78±6.09 |  |  | 29.68±9.00 |  |  | 2.69±1.36 |  |  | 5.61±1.90 |  |  |
| **COVID anyone around** |  |  |  |  |  |  |  |  |  |  |  |  |  |
| Yes | 387 (19.3) | 20.59±4.20 | -.856<sup>a</sup> | .392 | 22.97±7.25 | 7.796<sup>a</sup> | .001 | 2.37±.96 | - | .001 | 6.12±1.62 | 5.669<sup>a</sup> | .001 |
| No | 1614 (80.7) | 20.88±6.41 |  |  | 29.08±9.15 |  |  | 2.76±1.42 | 5.097<sup>a</sup> |  | 5.52±1.93 |  |  |

<sup>a</sup> Independent t test; <sup>b</sup> Chi-square test; \*

**Table 2:**
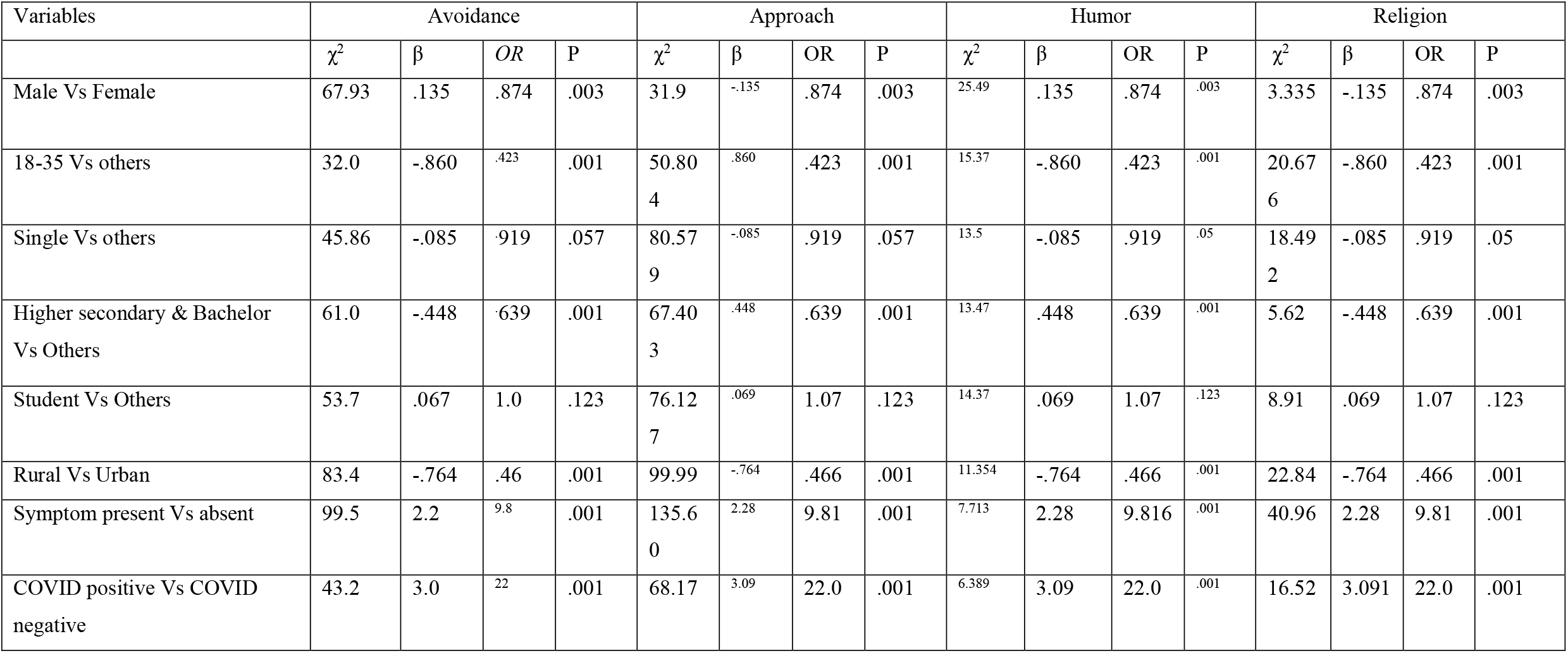
Binary Logistic regression of Socio-demographics and Coping strategy

## RESULTS

2001 participants aged 18 years to 86 years of age (31.85±14.2 years) responded to the survey. Response from male participants was 53.4% (n=1074) and female participants was 46.6% (n=927). Respondents represented all eight of the administrative divisions within Bangladesh, including: (1) Dhaka 63% (n=1261), (2) Chittagong 12.8% (n=258), (3) Rajshahi 11% (n=221), (4) Sylhet 04% (n=9), (5) Rangpur 4% (n=82), (6) Barishal 1.4% (n=29), (7) Khulna 3.4% (n=69), and (8) Mymenshing 3.5% (n=72). They also represent 55 out of 64 districts of Bangladesh. The respondent distribution is presented in Figure 1.

**Figure 1:**
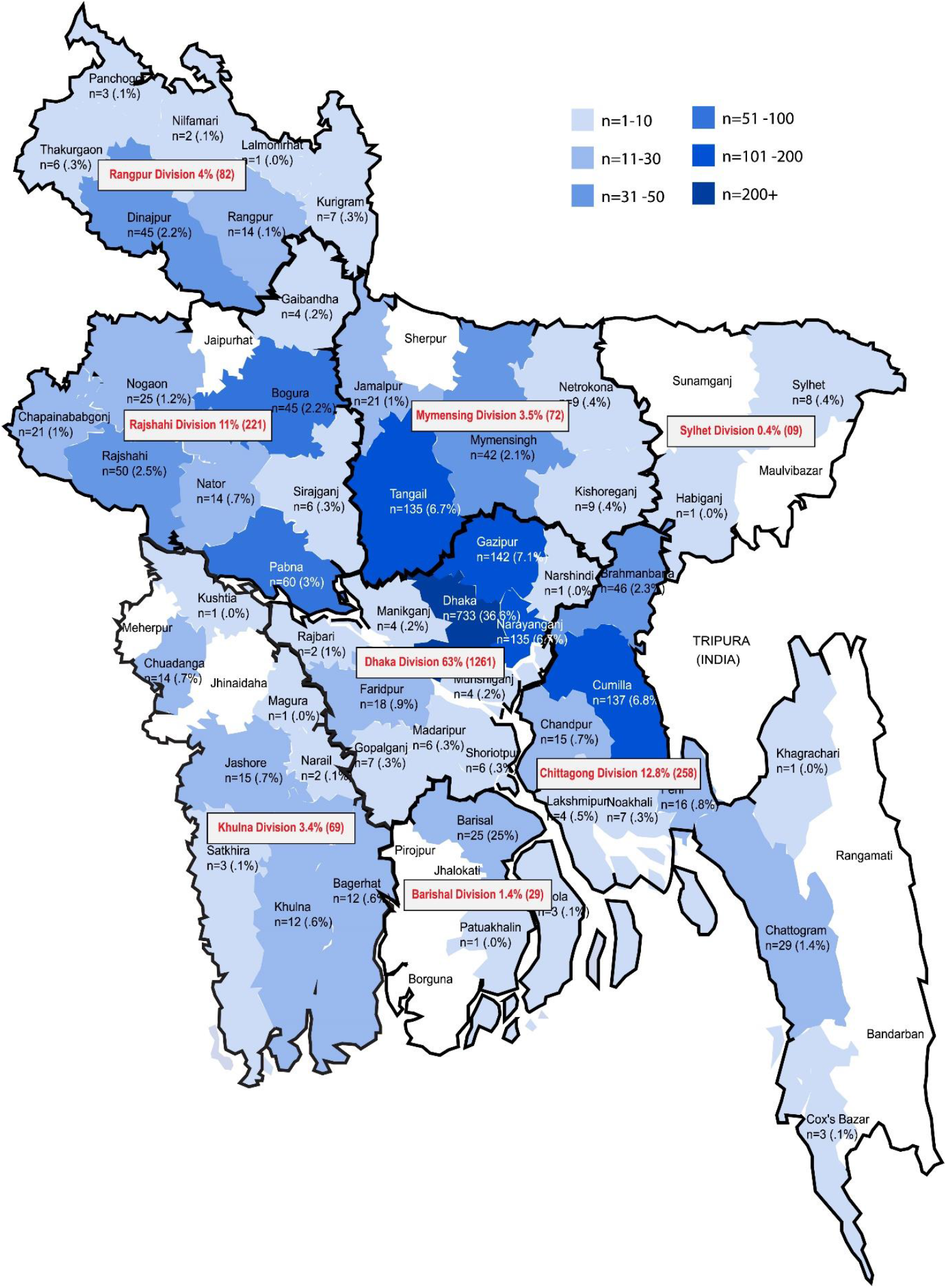
The population distribution of the respondents

The majority of the responses came from the younger population (18-35 years) 70.3% (n=1406) followed by middle aged adults (36-60 years) 24.8% (n=496). More than half (52.1%) were single or divorced, and 44.4% were married. The educational status varied from “no formal education” at 4% (n=81), and “primary education” at 9.5% (n=190) to “secondary education” 16.2% (n= 325), “higher secondary education” at 35.6% (n=713), bachelor’s degree at 25.4% (n=508), master’s degree at 8.5% (n=170) and Ph. D. degree reported by .7% (n=14) of respondents. In occupational variants, a major part of the respondents reported were students 48.3% (n=966); however, 17.9% (n=359) reported housewife as their major occupation; 9.9% (n=199) reported being employed in a private job; 5.8% (n=117) worked in a government job; 2.6% were retired; 1.7% (n=34) were unemployed; and 2.9% (n=59) reported other jobs. The “other jobs” reported related mostly to daily work. Approximately 68% (n=1365) of the respondents lived in the urban area, mostly near the district or sub-district of the Upozila Region and others were respondents from remote villages. In addition, 9.2% (n=185) reported COVID-like symptoms once or more in the period of March 2020 up to the data collection period. In multiple response analysis, COVID-19-like symptoms reported were similar to a fever with common cold symptoms, reported at 5.1% (n=103); fever, sore throat, cough with sputum were reported at 2.1% (n=42); viral symptoms with pneumonia were reported at 1% (n=20); viral symptoms with anosmia and sicca syndrome at .7% (n=14); and virus symptoms with diarrhea at .1% (n=3). 4.3% (n=87) were COVID positive in the RT-PCR test. Three hundred eight seven (n=387) respondents (19.3%) reported being exposed to someone close to them with a positive COVID-19 test. No known COVID-19 positive cases were interviewed during the data collection phase in maintenance of the study protocol. The detailed socio-demographics are presented in Table 1.

The Bangladeshi respondents showed a mixed coping strategy during the first wave of COVID-19. Higher scores were reported for approach coping strategies (APC) (29.83±8.9; Range: 12-48); avoidance coping strategies (AVC) were reported at lower levels, overall (20.83 ± 6.05; Range: 12-48). Humor (HU) scores were reported at 2.68±1.3 (2-8 scoring scale), and Religion (RE) scores were reported 5.64±1.8 (2 to 8 scoring scale). Figure 1 demonstrates that, among the respondents having the AVC style, 84.7% had a 2-3 score related specifically to mild to moderate substance use. The same group also reported similar scores for denial (72.4%), behavioral disengagement (61.4%) and self-blames (67.3%). Respondents with APC style had more scores of 6 to 8 (medium amount to all-time) related to inactive coping (45.3%), emotional support (37.2%), use of information support (35.5%), positive reframing (36.6%), planning (34%) and acceptance (55.1%). From the cluster sample, 24.5% of Bangladeshi respondents coped with the COVID-19 pandemic “all the time” and 30.8% coped “majority of the time” based on religious belief (RE) during the pandemic time frame for this study (Figure 2).

**Figure 2:**
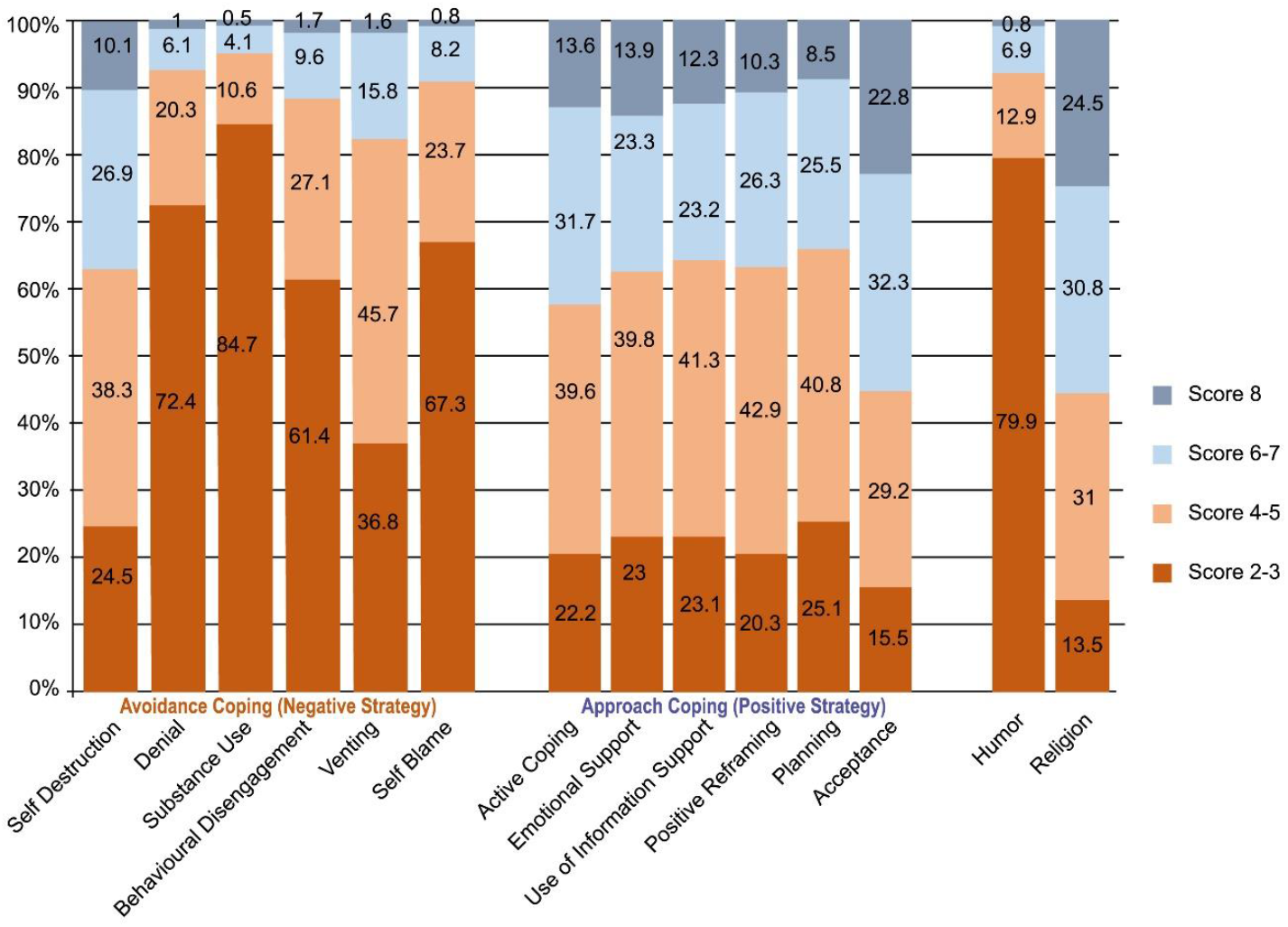
Population distribution of Coping

Male and females showed similar coping strategies, but male respondents reported higher score means for AVC (21.39±6.0, 20.18±5.9) and APC (30.13±8.7, 29.49±9.1) style in 12-48 scales compared to female respondents (Table 1). Male gender was associated with AVC (p<.01), and humor (p=<.01); whereas, no association was found between gender and APC style. Further multivariate analysis found a significant positive relationship between male gender and AVC style (χ2 67.9, β .135, OR.874, P <.01) and Humor (HU) coping style (χ2 25.4, β .135, OR.874, P <.01); inverse relationships were found between male gender and APC style (χ2 31.9, β-.135, OR.874, P <.01) and Religion (RE) coping style (χ2 3.335, β .135, OR.874, P <.01) (Table 2). In the Pearson Correlation Coefficient (PCC), overall age scores showed a statistically significant negative correlation with HU (r= -.06, P<.01) and RE (r= -.101, P<.01). Categorization of age group also found no statistically significant relationships with coping categories. However, respondents, aged 18-35 years, reported significantly higher scores related to AVC styles (χ2 32.0, β -.860, OR.42, P <.01), ACP styles (χ2 50.8, β .86, OR.42, P <.01), HU coping styles (χ2 15.37, β -.86, OR.42, P <.01), and RE coping styles (χ2 20.6, β -.86, OR.43, P <.01).

Marital status was positively associated with AVC styles (χ2 45.8, β -.085, OR.919, P <.01), approach (χ2 80.57, β -.085, OR.919, P <.01), HU (χ2 43.5, β -.085, OR.919, P <.01) and RE (χ2 18.492, β -.448, OR.69, P <.01). Similarly, Education had a statistically significant relationship (p <.01) with all categories of coping strategy. In binary logistic regression, Higher secondary and bachelor’s education groups reported an inverse relationship with AVC style (χ2 61, β -.448, OR.63, P <.01) and RE (χ2 5.62, β -.448, OR.69, P <.01), but exhibited a significant linear relationship with APC style (χ2 67.4, β .448, OR.63, P <.01) related to other education sub-categories. Occupation had a significant positive relationship with approach (p <.01). Rural geographical location was significantly different compared to urban locations in AVC style (χ2 83.4, β -.76, OR.46, P <.01), APC style (χ2 99.9, β -.76, OR.46, P <.01), HU (χ2 11.3, β -.76, OR.46, P <.01), and RE (χ2 22.8, β -.76, OR.46, P <.01). The respondents who had COVID-19 like symptoms had a relationship with AVC (P <.01), APC, and RE (P <.01). Respondents who experienced COVID-19 like symptom also showed a a significant difference with asymptomatic respondents with respect to AVC (P <.01). Respondents who tested COVID positive (n=87) significantly associated with APC (P <.01) and RE (P <.01) compared to respondents exposed to COVID positive cases in close contact who showed positive association with APC, HU, and RE (P <.01). Details are provided in Table 1 and Table 2.

In, Figure 3, the explanatory factor analysis revealed 2 major factors that were strongly associated with the coping items. A Kaiser-Meyer-Olkin (KMO) Test for Sampling Adequacy found .91 (superior to .50) and Bartlett’s Test of Sphericity was .001. Factor 1 found eigenvalue 5.645 (>1.14 and factor 2 found eigenvalue 3.010 (>1.14), other factors were not found eligible. In the principal component analysis using with a Varimax rotation and the minimum threshold for each factor to be considered at .40, the coping items found two clusters with a significant positive correlation (Figure 3). Factor 1 was associated with self-destruction (.739), venting (.670), active coping (.771), seeking emotional support (.800), seeking information support (.833), positive reframing (.753), planning (.759), acceptance (.556) and religion (.635). Factor 2 was associated with denial (.659), substance use (.716), behavior disengagement (.580), self-blame (.616) and humor (.752).

**Figure 3:**
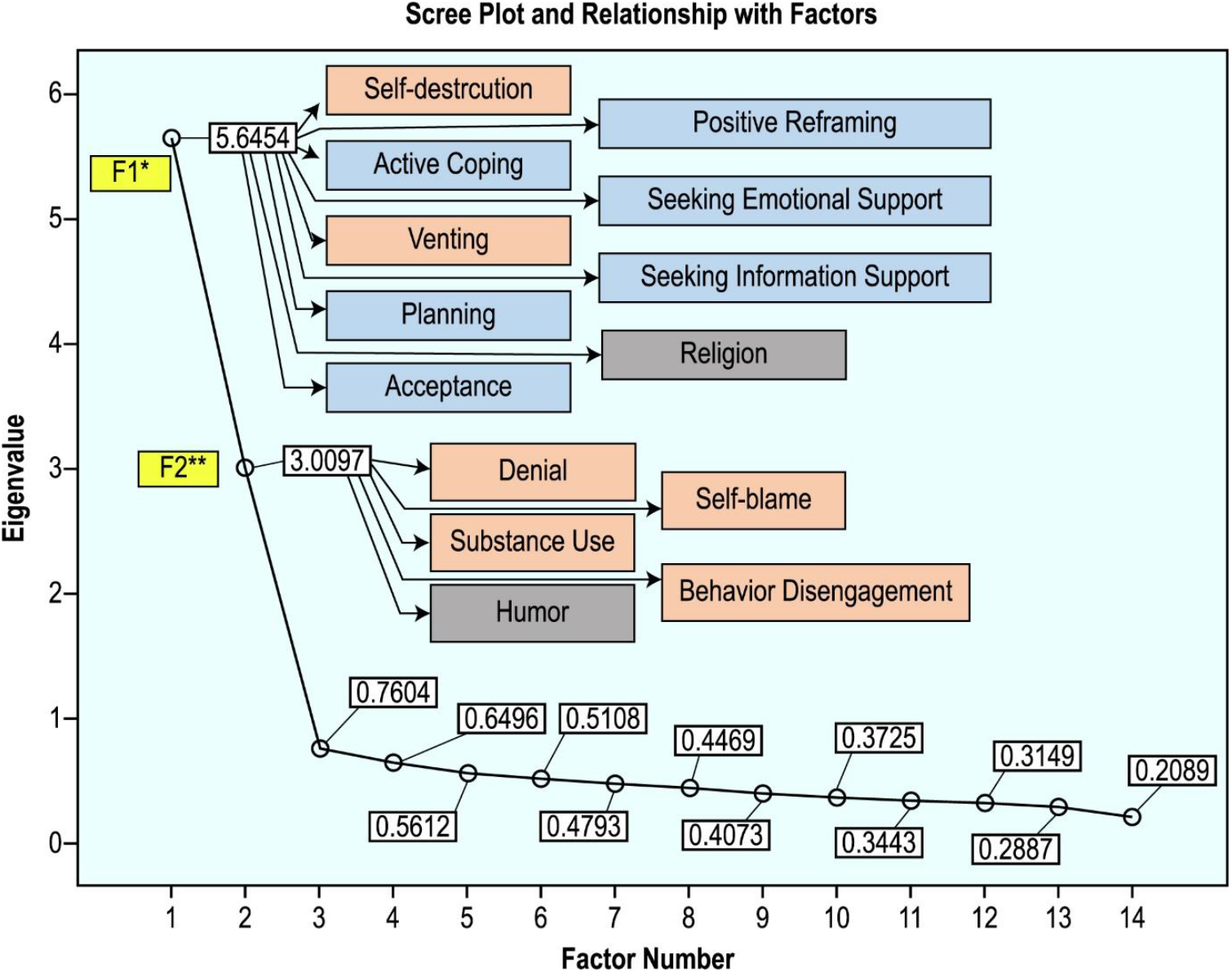
Exploratory Factor analysis of Categories of coping strategy * Factor 1 (eigenvalue 5.645 >1.1.4) was associated (Correlation matrix value superior to .40) with self-destruction (.739), venting (.670), active coping (.771), seeking emotional support (.800), seeking information support (.833), positive reframing (.753), planning (.759), acceptance (.556) and religion (.635). ** Factor 2 (eigenvalue 3.010 >1.1.4) was associated (Correlation matrix value superior to .40) with denial (.659), substance use (.716), behavior disengagement (.580), self-blame 477 (.616) and humor (.752).

## DISCUSSION

This community-based population study of coping strategies for COVID-19, derived from both rural and urban settings across Bangladesh and representing eight geographical divisions, found significantly higher prevalence of the Approach Coping Strategy (APC) compared to Avoidance Coping Strategy (AVC), overall. In addition, the overall reliance on Religion (RE) as a coping strategy was moderately strong compared to Humor (HU). To our best knowledge this is a groundbreaking population-based-survey on coping strategies utilized during the COVID-19 pandemic in Bangladesh. Results of this study reflect a mixed-approach coping style, with two major factors showing significant association with individual Brief-COPE domains.

In the population, higher scores (8 out of 8) were found to be more prevalent in APC and RE and lower scores (2-3 out of 8) were more prevalent in AVC and HU (Figure 2). Population mean scores showed a higher positive coping strategy, similar to APC (29.83±8.9). However, significant mean scores were also observed for negative coping strategies related to AVC, such as struggling with poor health status (20.83 ± 6.05). Higher scores on the APC were also related to higher scores, overall, on self-acceptance, active coping, emotional support, positive reframing, information support and planning. Furthermore, higher mean scores on APC strategies were positively associated with higher education levels, occupation, rural location, and positive COVID-19 status, positive PCR test result, and symptoms. AVC was associated with marital status, education less than secondary school, rural geographical location and having positive COVID-19 symptoms. AVC scores were significantly higher in males compared to females, showing higher dysfunctional coping strategies in Bangladeshi men. Bangladeshi respondents, both male and female, reported a significantly high score for RE coping (5.64±1.8 in the 2 to 8 score range). However, there were no significant differences by gender in reported scores by RE or APC styles.

Significantly higher mean scores were reported for the age group 18-35 compared to all age categories with respect to AVC, APC, HU, and RE. Marital status was significantly and strongly correlated with AVC, APC, HU, and RE, with mean scores higher for “single” status compared to all other categories. Religious coping has been found to be the most frequently utilized coping strategy for building resiliency in the face of life and health stress in Bangladesh. ^[42-43]^ It is not surprising, therefore, to find a similar high report of reliance on religion as one of the major coping strategies during the COVID-19 pandemic, for both men and women, living in various regions throughout Bangladesh. Religious coping strategy has not been categorized in either APC or AVC strategies and can be considered either a positive or negative coping strategy, depending on the perspective of the individual. Though this study did not focus on the specific positivity or negativity of religious coping strategies, there is evidence that religious coping strategies in Bangladesh are considered as positive support and are often mixed with traditional healing practices for over 80% of the population. ^[44]^ Older adults in our study who showed significant association to APC, similar to other studies, may be relying on resilience and coping mechanisms from previous life experiences. ^[25]^ Previous studies suggest, greater education ^[26]^ is linked to positive coping approaches; whereas female gender and single or separated people report more negative coping strategies resulting in poor health outcomes. ^[27]^ In our study, we found a high prevalence of alcohol (mild to moderate) use in male respondents as a means to cope for men and compared to very low prevalence of alcohol use in women for those who identified in the AVC style. A high prevalence of alcohol utilization for coping in our study is surprising, given that other studies have indicated 94.4% abstinence and only 5.6% lifetime prevalence in studies across Bangladesh. ^[45]^ In addition, women are much less likely to consume alcohol in Bangladesh because of socio-cultural norms, legal prohibition, and religious adherence expectations and when they do consume it, they may be much less likely to report use because of gendered norm expectations. ^[46]^ Humor, though significantly higher coping strategy in males compared to females, was not a major factor in coping for the overall population. The contributing factors behind higher scores of APC have not yet been examined. From our previous population-based survey ^[9]^ higher knowledge, education and female gender was correlated with positive attitude and practice towards COVID-19. The AVC approach was presumed to be higher, as one other study reported the seriousness of the mental health crisis during COVID-19 pandemic in Bangladesh. However, the results of our study show, overall, the APC style had higher overall scores among all participants. ^[10]^ AVC style was significantly higher in males compared to females, in our study, and has been shown to be directly linked to fear, ^[9]^ stress, ^[19]^ physical illness, insomnia, ^[20]^ misleading information or spreading of unreliable information, ^[21-23]^ that was not examined in this study. Differences, by gender, in coping styles may need further exploration to further evaluate all related variables. Exploratory factor analysis found a higher score of eigenvalues 5.645 and 3.010 (Cutoff 1.1.04) associated to 2 factors. The categories of coping strategies are highly associated in a positive direction to each other (correlation matrix value superior to .40). Factor 1 association was found for a mixed coping manner. The components of Factor 1 association included a mixture of AVC and APC styles involving: (1) self-destruction, (2) venting, (3) active coping, (4) seeking emotional support, (5) seeking information support, (6) positive reframing, (7) planning, (8) acceptance and (9) religion. Factor 2 was associated with a mixture of HU coping and AVC domains in denial, substance use, behavior disengagement, and self-blame. In a comparison study of a low-income nation, caregivers helping with HIV positive patients found 2 factors associated with APC and another 3 factors associated with mixed coping (APC, AVC, RE, and HU) in an exploratory factor analysis also using the Brief-COPE. ^[47]^ Our study sample represented 8 administrative divisions in Bangladesh, with only 2 divisions reporting less than one-third of the represented cluster sample size. We postulate that our study has adequate representation and strength and the results may be externally generalized to all of Bangladesh. Our study also contributes to the understanding of predominant coping strategies utilized by the Bangladeshi people during the COVID-19 pandemic and may, potentially, contribute to positive health policies related to the promotion of mental health for Bangladeshi citizens in the future.

One limitation of our study is that the Brief-COPE is a self-report measurement tool and, though highly valid and reliable for major domain areas such as Avoidance, Approach, Humor and Religion, some of the individual subscales show lower internal consistency, when considered in a separate analysis. However, for this sample and the report of the four domains avoidance, approach, humor and religion the validity of the instrument is considered reasonable and acceptable. This study also overcame the barriers to extensive data collection within a short period of time by training skilled rehabilitation data collectors, who lived in the local regions to conduct the face-to-face interviews for all participating respondents. Another limitation of the study is the lack of pre-pandemic coping strategy data to compare the overall effect of COVID-19 on coping strategy changes over time. Furthermore, examination of specific detailed strategies of coping, particularly related to religious coping, would further clarify meaning, both positive and negative strategies, in the context of a major infectious disease threat. Future research would be improved by gaining more understanding of the concept of resilience, while including consideration of how people with disability, ^[48]^ refugees, ^[49]^ migrant workers, and other marginalized populations cope with adversities caused by the pandemic. This research could also include the role of traditional and local healing medicine as they relate to religious coping strategies in different regions. Future research could also explore coping strategies and health equity concerns related to vulnerable populations across the region and internationally to understand coping strategies in a global context. Resilience and coping could then be explored in combination with other variables such as region, race, gender identity, disability status and legal status in the case of people with migrant and refugee status.

## CONCLUSION

Two cluster factors were identified as significant coping style groups for the respondents in this study. Factor 1 was identified as a mixture of AVC and APC styles, including self-destruction, venting, active coping, seeking emotional support, seeking information support, positive reframing, planning, acceptance and religion. Factor 2 was identified as a mixture of AVC and HU coping styles. Overall, a higher utilization of APC was associated with better responses to adverse circumstances, better physical and mental health outcomes. Male respondents reported utilization of more coping strategies in each category (APC, AVC, & HU), except religion, than females. Within the AVC strategy, alcohol utilization and denial showed higher than expected prevalence rates in men. Religion was found to be an integral component of coping all respondents. Limitations include study design and lack of detailed information regarding positivity or negativity the RE and HU strategies reported. Future research into the concept of resilience and how this could be of practical benefit to individuals, families and communities would be extremely useful given that COVID-19 is here to stay. It could also be useful to examine the differences in coping strategies for vulnerable populations and consider any potential differences detected in relation to region, race, gender identity, disability status, legal status in the case of people with migrant and refugee status.

## Data Availability

The data are available regarding this study and can be viewed in https://www.kaggle.com/kmamranhossain/population-cope

## ACKNOWLEDGEMENT

Authors acknowledge the students of Bangladesh Health Professions Institute (BHPI) for their voluntary contribution in data collection.

## SUPPORTING INFORMATION

S1 File: Questionnaire English and Bangla

S2 File: Helsinki Declaration

S1 Table: Socio-demographic distribution and relationship with the coping domain

S2 Table: Binary Logistic regression of Socio-demographics and Coping strategy

S1 Figure: The population distribution of the respondents

S2 Figure: Population distribution of Coping

S3 Figure: Exploratory Factor analysis of Categories of coping strategy

